# Monitoring for 5-aminosalicylate toxicity: prognostic model development and validation

**DOI:** 10.1101/2023.12.15.23299944

**Authors:** A Abhishek, Georgina Nakafero, Matthew J Grainge, Tim Card, Maarten W Taal, Guruprasad P Aithal, Christopher P Fox, Christian D Mallen, Matthew D Stevenson, Richard D Riley

**Affiliations:** School of Medicine, University of Nottingham, Nottingham NG5 1PB, UK; Lifespan and Population Health, School of Medicine, University of Nottingham, Nottingham NG5 1PB, UK; Centre for Kidney Research and Innovation, Translational Medical Sciences, School of Medicine, University of Nottingham, Derby DE22 3NE, UK; Nottingham Digestive Diseases Centre, Translational Medical Sciences, School of Medicine, University of Nottingham, Nottingham NG7 2UH, UK; Department of Haematology, Nottingham University Hospital NHS Trust, Nottingham NG5 1PB, UK; Primary Care Centre Versus Arthritis, Keele University, Keele ST5 5BJ, UK; School of Health and Related Research, University of Sheffield, Sheffield S1 4DA; Institute of Applied Health Research, College of Medical and Dental Sciences, University of Birmingham, Birmingham B15 2TT, UK

**Author notes:** **Corresponding author:** Prof. Abhishek. **Address for correspondence:** A23, City Hospital Nottingham, The University of Nottingham, Nottingham NG5 1PB, UK**. Email:**.

**Keywords:** 5-aminosalicylate, nephritis, hepatotoxicity, myelotoxicity

## Abstract

**Background and aim:** To develop and validate a prognostic model for risk-stratified monitoring of 5-aminosalicylate (5-ASA) toxicity.

**Methods:** This nationwide retrospective cohort study used data from the Clinical Practice Research Datalink (CPRD) Aurum and Gold for model development and validation respectively. It included adults newly diagnosed with inflammatory bowel disease (IBD) and established on 5-ASAs between 01/01/2007 and 31/12/2019. 5-ASA discontinuation with abnormal monitoring blood test result was the outcome of interest. Patients prescribed 5-ASAs for ≥6 months i.e., established on treatment, were followed-up for up to five years. Penalised Cox-regression was used to develop the risk equation. Model performance was assessed in terms of calibration and discrimination. Statistical analysis was performed using STATA (StataCorp LLC).

**Results:** 14,109 and 7,523 participants formed the development and validation cohorts with 401 and 243 events respectively. 185, 172, and 64 discontinuations were due to cytopenia, elevated creatinine and elevated liver enzymes respectively in the derivation cohort. Hazardous alcohol intake, chronic kidney disease, thiopurine use, and blood test abnormalities before follow-up were strong prognostic factors. The optimism adjusted R^2^_D_ in development data was 0.08. The calibration slope and Royston D statistic (95% Confidence Interval) in validation cohort were 0.90 (0.61-1.19) and 0.57 (0.37-0.77) respectively.

**Conclusion:** This prognostic model utilises information available during routine clinical care and can be used to inform decisions on the interval between monitoring blood-tests. The results of this study ought to be considered by guideline writing groups to risk-stratify blood test monitoring during established 5-ASA treatment.

**What is already known?:**

- Renal, hepatic and blood toxicity are uncommon during long-term 5-aminosalicylate (5-ASA) treatment.
- There are no mechanisms to predict the risk of these toxicities during established treatment that may be used to risk stratify blood-test monitoring.

**What this study adds?:**

- Using a large national dataset originated during routine care, this study developed a prognostic model that discriminated patients at varying risk of 5-ASA toxicity during established treatment with good performance characteristics validated.
- Most patients were at low-risk of toxicity due to 5-ASAs and could continue with annual monitoring blood-tests while others at high risk may require more frequent monitoring.
- This prognostic model can be used to make an informed decision on the interval between monitoring blood tests and the findings ought to be considered by guideline writing groups to bring about equitable and sustainable change in clinical practice.

## Introduction

5-aminosalicylates (5-ASA) are the mainstay of treatment for mild to moderate ulcerative colitis and are often combined with biologics in the treatment of severe disease^1–3^. Although effective, they can cause side-effects such as interstitial nephritis, hepatitis and blood-dyscrasias^4–20^. These side-effects are common early during the treatment^17–19^ but they also occur later. For instance, the median time to development of interstitial nephritis ranged between 2.3 and 3 years in published studies^8,9^. There is substantial variation in recommendations on screening for the occurrence of myelotoxicity, hepatotoxicity, and nephrotoxicity during established 5-ASA treatment.

For instance, the summary of product characteristics recommends three monthly full blood count (FBC), liver function test (LFT), and urea electrolytes and creatinine (UE&C) during established treatment^19^, whereas the European consensus from the European Crohn’s and Colitis Organisation is to monitor with FBC and UE&C at three to six monthly intervals^3^, while the American Gastroenterology Association and the British Society of Gastroenterology recommend monitoring with UE&C periodically and annually respectively during established treatment^1,2^. This latter may be disadvantageous to patients treated with 5-ASA drugs due to their well-documented but uncommon myelotoxicity and hepatotoxicity.

Due to the rarity of cytopenia, hepatotoxicity and nephrotoxicity due to 5-ASA^4–6,16^, It would be beneficial to predict clinically significant renal, hepatic or myelotoxicity during established treatment to allow risk-stratified monitoring. This study developed and validated a prognostic model for clinically significant 5-ASA toxicity during established treatment.

## Methods

### Data source

Data from the Clinical Practice Research Datalink (CPRD) Aurum and Gold were used for model development and validation respectively ^21,22^. CPRD is an anonymised longitudinal database of electronic health records collected during routine clinical care in the NHS. With almost universal coverage of UK residents, participants that contributed data to the CPRD are representative of the UK population ^21^. CPRD Aurum and Gold complement each other in terms of coverage of general practices according to the use of different software for data capture. Some general practices that have contributed data to both databases were only included in the model development cohort.

### Approvals

ISAC of the MHRA (Reference: 20_000236R).

### Study design

Retrospective cohort study.

### Study period

1^st^ January 2007 to 31^st^ December 2019.

### Study population

Participants aged ≥18 years with a new diagnosis of inflammatory bowel disease (IBD) newly prescribed 5-ASA by their GP for at-least six-months were eligible (Supplementary methods). Patients with severe liver, kidney or haematological diseases prior to first 5-ASA prescription were excluded as described previously^23^.

### Follow-up

Patients were followed-up from 180 days after their first GP prescription until the earliest of outcome, death, transfer out of practice, 90-days prescription gap, last data collection from practice, 31/12/2019 or five-years from the start of follow-up.

### Outcome

5-ASA-toxicity associated drug discontinuation was the outcome of interest. This was defined as a prescription gap of ≥90 days with either an abnormal blood-test result (either leucocyte count <3.5×10^9^/L, or neutrophil count <1.6×10^9^/L, or platelet count <140×10^9^/L, or alanine transaminase and/or aspartate transaminase >100 IU/mL, or decline in kidney function, defined as either progression of chronic kidney disease (CKD) based on medical codes recorded by the GP, or >26 μmol/L increase in creatinine concentration, the threshold for consideration of acute kidney injury (AKI)^24^) or a diagnostic code for abnormal blood-test result within ±60 days of the last prescription date^23,25^.

A random sample of 5-ASA discontinuations with abnormal blood test results was drawn. Data for all diagnostic codes entered during primary-care consultations within ±60 days of the abnormal blood test result were extracted. A.A. screened the list to identify outcomes that could potentially be explained by an alternative condition or its treatment.

*<u>Predictors</u>* were selected based on clinical expertise and knowledge of the published literature (Table 1). Age, sex, body mass index (BMI), alcohol intake, and diabetes were included as they associate with drug induced liver injury (DILI) ^26,27^. CKD was included as it reduces 5-ASA clearance^28^. Statins, carbamazepine, valproate, paracetamol were included as their use is associated with 5-ASA toxicity relevant to this study^28^. Methotrexate, leflunomide, thiopurines were included as they can cause cytopenia, elevated liver enzymes and AKI. Either cytopenia (neutrophil count <2 x 10^9^/l, or total leucocyte count <4 x 10^9^/l, or platelet count <150 10^9^/l) or elevated transaminase (ALT and/or AST >35 IU/l) during the first six months of primary care prescription were included as they predicted cytopenia and/or transaminitis in other studies ^29,30^.

**Table 1:** Distribution of candidate predictors in development and validation cohorts

| Predictor<sup>1</sup> | Derivation cohort<br>(CPRD Aurum)<br>n=14,109 | Validation cohort<br>(CPRD Gold)<br>n=7,523 |
| --- | --- | --- |
| Age, mean (standard deviation) year | 46.7 (17.6) | 47.5 (17.7) |
| Male sex | 7,253 (51.4) | 3,866 (51.4) |
| Body mass index |  |  |
| <18.5 kg/m<sup>2</sup> | 436 (3.1) | 234 (3.1) |
| 18.5-24.9 kg/m<sup>2</sup> | 5,033 (35.7) | 2,540 (33.8) |
| 25.0-29.9 kg/m<sup>2</sup> | 4,047 (28.7) | 2,247 (29.9) |
| ≥30 kg/m<sup>2</sup> | 2,660 (18.7) | 1,482 (19.7) |
| Missing | 1,933 (13.7) | 1,020 (13.6) |
| Alcohol use |  |  |
| Non-user | 2,373 (16.8) | 936 (12.4) |
| Low (1-14 units/week) | 5,838 (41.4) | 3,815 (50.7) |
| Moderate (15-21 units/week) | 847 (6.0) | 429 (5.7) |
| Hazardous (>21 units/week) | 1,024 (7.3) | 419 (5.6) |
| Ex-user | 1,075 (7.6) | 400 (5.3) |
| Missing | 2,952 (20.9) | 1,524 (20.3) |
| Inflammatory conditions |  |  |
| Inflammatory bowel disease alone | 13,728 (97.3) | 7,318 (97.3) |
| Psoriasis | 276 (2.0) | 141 (1.9) |
| Autoimmune rheumatic disease<sup>2</sup> | 105 (0.7) | 64 (0.9) |
| Comorbidities |  |  |
| Diabetes | 1,099 (7.8) | 582 (7.7) |
| Chronic kidney disease stage-3 | 827 (5.9) | 4.8 (5.4) |
| Immunosuppressive drugs |  |  |
| Thiopurine | 1,336 (9.5) | 814 (10.8) |
| Methotrexate/leflunomide | 64 (0.5) | 24 (0.3) |
| Other drugs |  |  |
| Statins | 2,078 (14.7) | 1,124 (14.5) |
| Carbamazepine/valproate | 119 (0.8) | 74 (1.0) |
| Paracetamol | 1,001 (7.1) | 653 (8.7) |
| At least mild cytopenia or liver<br>enzyme elevation in six-months<br>preceding start of follow-up | 973 (6.9) | 516 (6.9) |

<sup>1</sup>Values are numbers (percentage) unless stated otherwise. <sup>2</sup>Rheumatoid arthritis, systemic lupus erythematosus, ankylosing spondylitis/Reactive arthritis. CPRD: Clinical Practice Research Datalink.

### Sample size

The incidence of toxicity from 5-ASA ranged between 1.2 and 1.7 per 1,000 person-years for interstitial nephritis^5,6^. To minimise model overfitting and ensure precise estimation of overall risk, the minimum sample size required for new model development was 2,590 participants (39 events) based on a maximum of 20 parameters, Cox-Snell R^2^ value of 0.12, estimated event rate of 0.005/person-year, a 5-year time horizon, and a mean follow-up period of 3 years using the formulae of Riley et al.,^31^. The sample size for external model validation was larger than the typically recommended minimum sample size of 200 events^32^.

### Statistical analysis

Multiple imputation handled missing data on BMI and alcohol intake using chained equations ^33^. We carried out 10 imputations in the development dataset and 5 imputations in the validation dataset - a pragmatic approach considering the large size of CPRD. The imputation model included all candidate predictors, baseline Nelson-Aalen cumulative hazard function and outcome variable.

### Model development

Fraction polynomial regression (first degree) analysis was used to model non-linear risk relationships with continuous predictors, but these were not better than the linear terms (p > 0.05), hence were not transformed. All candidate predictors and parameters were included in the Cox model and coefficients of each parameter estimated and combined using Rubin’s rule across the imputed datasets. The risk equation for predicting an individual’s risk of 5-ASA discontinuation with abnormal blood-test results by 5-years follow-up was formulated in the development data. The baseline survival function at *t=5 years*, a non-parametric estimate of survival function when all predictor values are set to zero, which is equivalent to the Kaplan-Meier product-limit estimate, was estimated along with the estimated regression coefficients (β) and the individual’s predictor values (X). This led to the equation for the predicted absolute risk over time ^34^:

Predicted risk = 1 – S_0_(t_=5_)^exp(Xβ)^ where S_0_(t_=5_) is the baseline survival function at 5-years of follow-up and βX is the linear predictor, β_1_x_1_+ β_2_x_2_+ … + β_p_x_p_.

### Model internal validation and shrinkage

The performance of the model in terms of calibration was assessed by plotting agreement between predicted and observed outcomes. Internal validation was performed to correct performance estimates for optimism due to overfitting by bootstrapping with replacement 500 samples of the development data. The full model was fitted in each bootstrap sample and then its performance was quantified in the bootstrap sample (apparent performance) and the original sample (test model performance), and the optimism calculated (difference in test performance and apparent performance). A uniform shrinkage factor was estimated as the average of calibration slopes from the bootstrap samples. This process was repeated for all 10 imputed datasets, and the final uniform shrinkage calculated by averaging across the estimated shrinkage estimates from each imputation. Optimism-adjusted estimates of performance for the original model were then calculated, as the original apparent performance minus the optimism.

To account for overfitting, the original β coefficients were multiplied by the final uniform shrinkage factor and the baseline hazards re-estimated conditional on the shrunken β coefficients to ensure that overall calibration was maintained, producing a final model. The D statistic, a measure of discrimination, interpreted as a log hazard ratio (HR) ^35,36^, and R^2^, a measure of variation explained by the model were calculated.

### Model external validation

The final developed model equation was applied to the validation dataset, and calibration and discrimination were examined^35,36^. Calibration of 5-year risks was examined by plotting agreement between estimated risk from the model and observed outcome risks. In the calibration plot, predicted and observed risks were divided into 10 equally sized groups. Pseudo-observations were used to construct smooth calibration curves across all individuals via a running non-parametric smoother. Separate graphs were plotted for each imputation. Stata-MP version 16 was used for statistical analyses. This study was reported in line with the TRIPOD guidelines ^37^.

## Results

### Participants

Data for 14,109 and 7,523 participants that contributed 41,146 and 21,070 person-years follow-up were included in the development and validation cohorts, respectively (Figure S1 and S2). Over 97% participants were diagnosed with IBD alone, over 50% self-identified as male, and had similar lifestyle factors, comorbidities, and drug treatments (Table 1). Twelve candidate predictors (17 predictor parameters) were included in the model (Table 2).

**Table 2:**
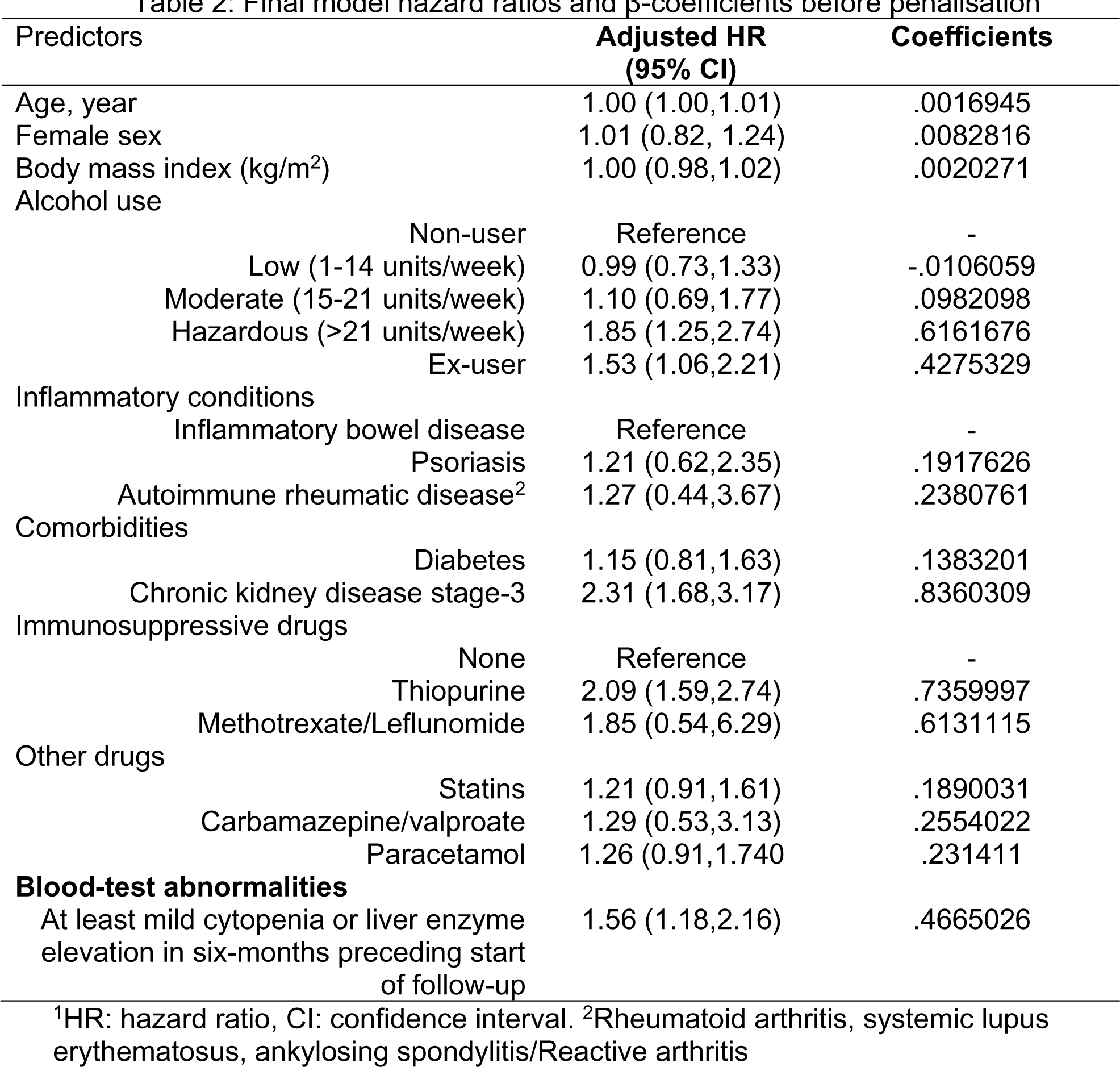
Final model hazard ratios and β-coefficients before penalisation

| Predictors | Adjusted HR<br>(95% CI) | Coefficients |
| --- | --- | --- |
| Age, year | 1.00 (1.00,1.01) | .0016945 |
| Female sex | 1.01 (0.82, 1.24) | .0082816 |
| Body mass index (kg/m<sup>2</sup>) | 1.00 (0.98,1.02) | .0020271 |
| Alcohol use |  |  |
| Non-user | Reference | - |
| Low (1-14 units/week) | 0.99 (0.73,1.33) | -.0106059 |
| Moderate (15-21 units/week) | 1.10 (0.69,1.77) | .0982098 |
| Hazardous (>21 units/week) | 1.85 (1.25,2.74) | .6161676 |
| Ex-user | 1.53 (1.06,2.21) | .4275329 |
| Inflammatory conditions |  |  |
| Inflammatory bowel disease | Reference | - |
| Psoriasis | 1.21 (0.62,2.35) | .1917626 |
| Autoimmune rheumatic disease<sup>2</sup> | 1.27 (0.44,3.67) | .2380761 |
| Comorbidities |  |  |
| Diabetes | 1.15 (0.81,1.63) | .1383201 |
| Chronic kidney disease stage-3 | 2.31 (1.68,3.17) | .8360309 |
| Immunosuppressive drugs |  |  |
| None | Reference | - |
| Thiopurine | 2.09 (1.59,2.74) | .7359997 |
| Methotrexate/Leflunomide | 1.85 (0.54,6.29) | .6131115 |
| Other drugs |  |  |
| Statins | 1.21 (0.91,1.61) | .1890031 |
| Carbamazepine/valproate | 1.29 (0.53,3.13) | .2554022 |
| Paracetamol | 1.26 (0.91,1.740) | .231411 |
| **Blood-test abnormalities** |  |  |
| At least mild cytopenia or liver enzyme elevation in six-months preceding start of follow-up | 1.56 (1.18,2.16) | .4665026 |

<sup>1</sup>HR: hazard ratio, CI: confidence interval. <sup>2</sup>Rheumatoid arthritis, systemic lupus erythematosus, ankylosing spondylitis/Reactive arthritis

### Model development

In the derivation dataset, 401 outcomes occurred in 2.8% patients (n=14,109) during follow-up at a rate (95% CI) of 9.75 (8.84 – 10.75) per 1,000 person-years. Outcome validation exercise in 167 outcomes revealed that only 10.2% outcomes (n=17) could potentially be explained by another contemporaneous illness or its treatments, with a positive predictive value of 89.8% (Table S1). Cytopenia, increase in serum creatinine, and elevated liver enzymes caused 5-ASA discontinuation in 185, 172, and 64 participants respectively (Figure S3). CKD-3, thiopurine co-prescription, hazardous alcohol use, ex-alcohol use and either cytopenia or elevated liver enzymes during first six months of 5-ASA prescription were strong predictors of drug discontinuation with adjusted hazard ratio (95% confidence interval (CI)) 2.31 (1.68,3.17), 2.09 (1.59,2.74), 1.85 (1.25,2.74), 1.53 (1.06,2.21) and 1.56 (1.18,2.16) respectively (Table 2). Before shrinkage, the calibration slope in the derivation data was 1.00 (95% CI 0.82-1.18). From the bootstrap, a uniform shrinkage factor of 0.83 was obtained and used to shrink predictor coefficients in the final model for optimism and after re-estimation, the final model’s cumulative baseline survival function (S_0_) was 0.971 at 5-years of follow-up (Box 1).

### Model performance in the derivation cohort

Calibration plot of the final (i.e., after shrinkage) model at 5-years showed that the average model predictions matched the average observed outcome probabilities across 10 groups of patients, with confidence intervals overlapping the 45-degree line (perfect prediction line) (Figure 1). As most patients had low risk of outcome, most of the deciles clustered at the bottom left of the calibration plot (Figure S4). The smoothed calibration curve at 5-years showed alignment of observed risk to the predicted risk with wide confidence intervals at >0.1 risk probabilities (Figure 1, Figure S5). Royston *D* statistic (95% CI) was 0.75 (0.59,0.91) corresponding to HR 2.12 (1.80-2.48) when comparing the risk groups above and below the median of linear predictor. The optimism adjusted Royston *D* statistic was 0.60 corresponding to HR 1.82, obtained by exponentiating the D statistic (Table 3).

**Figure 1:**
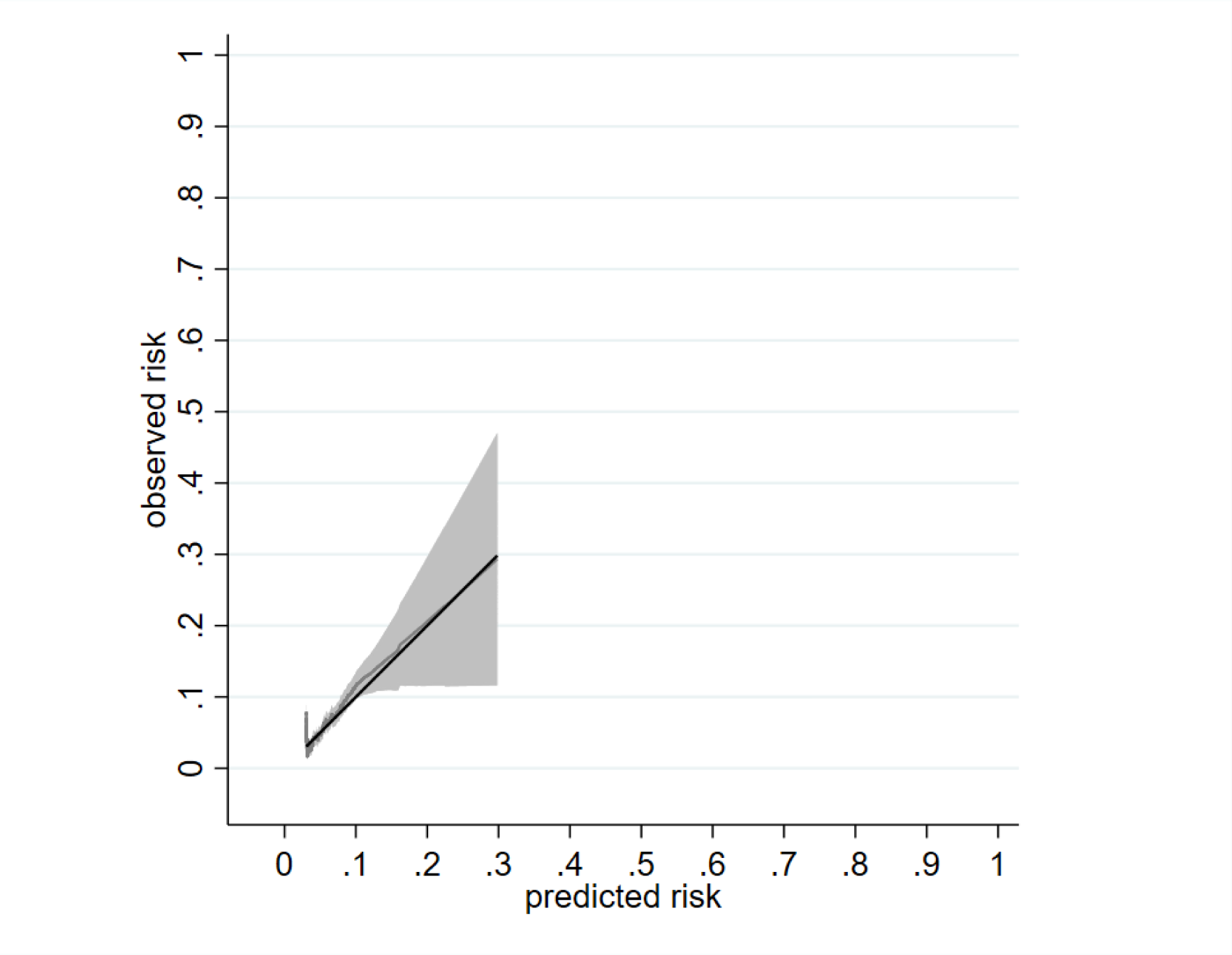
Calibration of a prognostic model for 5-ASA discontinuation with abnormal monitoring blood-test results at 5 years in the development cohort^1^. ^1^Data from a single imputed dataset was used; S_o_(t_=5_) =0.971

**Table 3:**
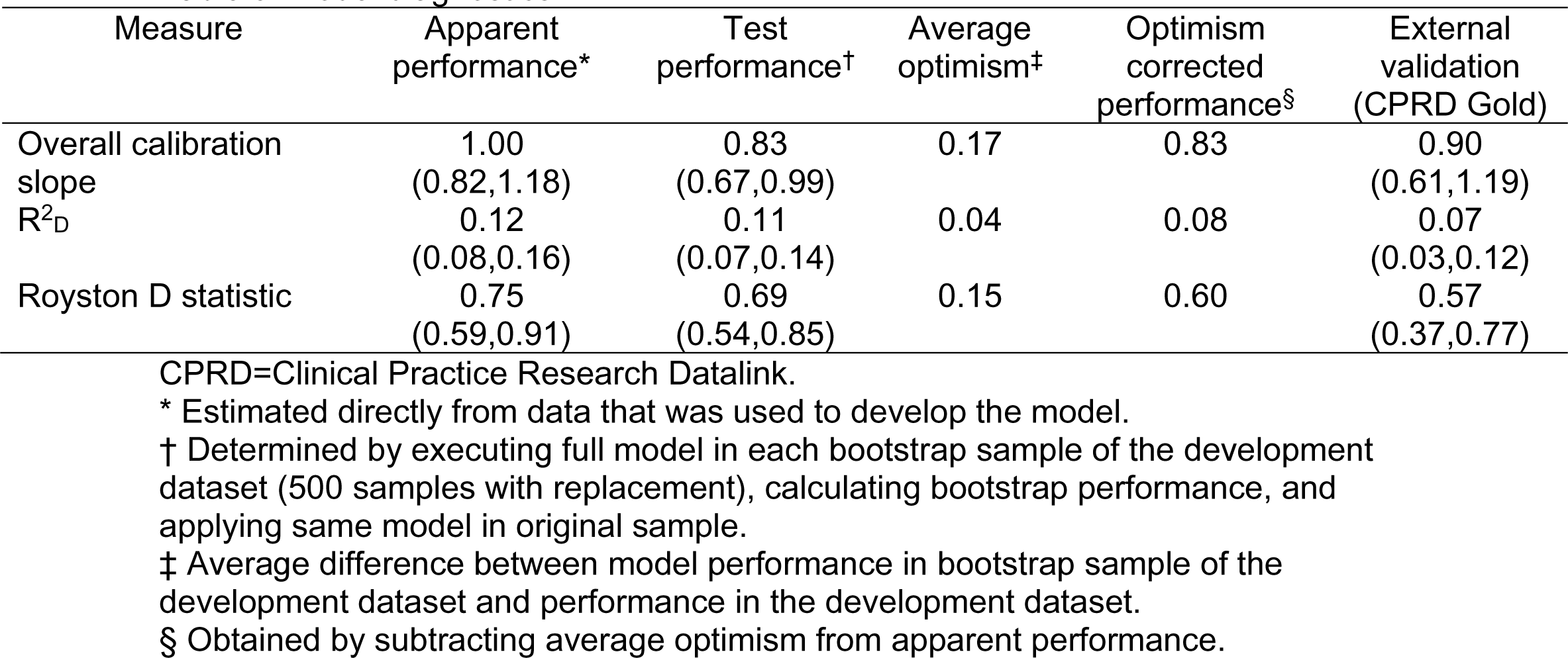
Model diagnostics

### Model performance in the validation cohort

There were 243 outcomes in 3.2% (n=7,523) patients at a rate (95% CI) of 11.53 (10.17-13.08) per 1000 person-years in the validation cohort. The calibration slope (95% CI) across the five-year follow-up period was 0.90 (0.61-1.19). The calibration plot showed reasonable correspondence between observed and predicted risk at 5-years across the tenths of risk with confidence intervals crossing the perfect prediction line (Figure S6). Most groups clustered at the bottom left of the calibration plot owing to low risk of outcome for most patients (Figures S7). When individual risk was plotted, the smoothed calibration curve showed instability in the model predictions with a tendency to under-predict low risk by a small amount (Figure 2). Model performance was also tested at years 1, 2, 3 and 4 and showed a similar pattern (Figures S8-11). Model discrimination in the derivation and validation data was broadly similar (Table 3). The Royston D statistic in the validation data was 0.57 (0.37,0.77), corresponding to HR (95% CI) 1.77 (1.45-2.16). The R^2^ score was 0.07 (95% CI 0.03,0.12).

**Figure 2:**
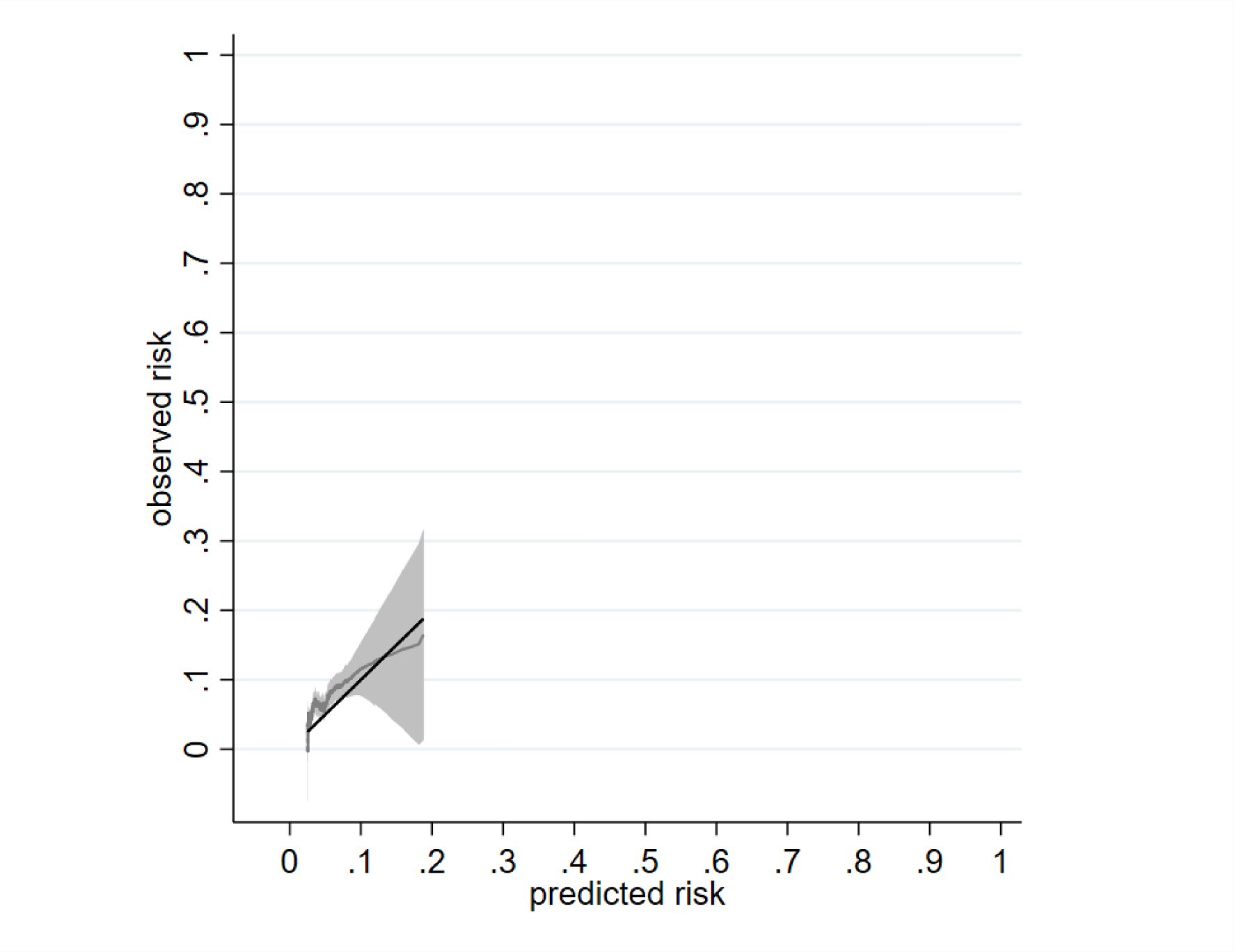
Calibration of a prognostic model for 5-ASA discontinuation with abnormal monitoring blood-test results at 5 years in the validation cohort^1^. ^1^Data from a single imputed dataset was used; So(t=5) =0.971

### Worked examples

Ten anonymised patient profiles, one from the middle of each decile of predicted risk were selected from the development cohort (Table S2). The cumulative probability of outcome over five years ranged from 3.1% in the middle of the first group to 4.6% in the middle of the seventh group, and 8.6% in the middle of the 10th group.

## Discussion

This study developed and externally validated a prognostic model for 5-ASA discontinuation with abnormal monitoring blood test results. To the best of our knowledge this is the first such endeavour. The model was developed using a large real-world and nationally representative dataset that originated during routine care of patients and has high generalisability. It had good performance characteristics in an independent dataset. It predicted clinically relevant toxicity i.e., one that required 5-ASA treatment being stopped, indicating face validity. The prognostic factors were selected to include those that are readily available during routine consultations. This makes the model is easy to use.

5-ASAs were stopped for cytopenia, elevated creatinine and elevated liver enzymes suggesting that monitoring blood-tests should include FBC, UE&C, and LFT. Numerically more patients discontinued 5-ASA due to cytopenia than due to nephrotoxicity, with far fewer participants stopping treatment due to elevated liver enzymes. In another previous study, there were numerically greater number of blood-dyscrasias than AKI with relatively few instances of elevated liver enzymes^4^. Similarly, a French pharmacovigilance study in which spontaneously reported adverse events were included and causality assessment was performed reported numerically more cases with myelotoxicity (n=7) and either hepatitis or elevated liver enzymes (n=7) than renal insufficiency (n=2) over a one-year period^16^. 5-ASA drugs are believed to be less hepatotoxic than sulfasalazine, however, the cumulative occurrence of drug induced liver injury was similar in the mesalazine and sulfasalazine arms of a randomised controlled trial, with 2.6% of patients taking mesalazine experiencing liver injury^20^. In another study, the mean white blood cell count reduced significantly with 5-ASA treatment, while the mean levels of creatinine and liver enzymes remained unchanged^15^. Thus, the findings of our study are consistent with previous reports.

CKD-3, hazardous and prior-alcohol use were strong independent predictors of 5-ASA discontinuation with blood-test abnormalities. These are novel findings. Nevertheless, these are plausible predictors. Co-prescription of a thiopurine was a strong risk-factor for toxicity consistent with other studies^38–40^. Abnormal blood-test results during the first six months of therapy were strong independent predictors of discontinuing 5-ASA with abnormal monitoring blood-test results, like previous reports about other drugs^23,41^.

However, several limitations of this study ought to be considered. First, information about the date when 5-ASA was first prescribed from hospital was unavailable. This is not a serious issue as the model can be applied once the patient has completed six-months of 5-ASA treatment from primary-care after treatment initiation and initial stabilization in a hospital clinic, or approximately one year from first prescription in a healthcare system without shared care prescription. Second, we did not have information on the use of biologics as these are hospital prescribed. Third, data on disease activity are not recorded in the CPRD. Fourth, we could not consider ulcerative colitis and Crohn’s disease as separate prognostic factors as it is not possible to reliably differentiate between them in the CPRD. Fifth, the abnormal blood test could be due to a different illness. Although in our previous validation studies, only 5.4% of abnormal blood-test results could be explained by an alternative illness ^25^. Sixth, although the external validation dataset was distinct from the model development dataset, it also originated from UK. We recommend that our model be validated in a dataset from another country. Seventh, there were 9 (0.06%) patients in the highest three tenths of risk resulting in uncertainty regarding predictors for these groups. Eighth, we did not perform competing risk regression. However, this does not limit the validity of our findings as there were few deaths (26 [0.18%]) in the derivation and (10 [0.13%]) validation cohorts up to the 5-year follow-up.

In conclusion, we have developed and externally validated a prognostic model for 5-ASA discontinuation with abnormal monitoring blood test results that may be used to individualise monitoring using principles of shared decision making between the patient and the physician. These findings need to be considered by guideline writing groups to bring about sustained change in clinical practice.

## Author contributions

A Abhishek: the conception and design of the study, interpretation of data, drafting the article, revising article critically for important intellectual content, final approval of the version to be submitted.

Georgina Nakafero: the conception and design of the study, analysis and interpretation of data, assisted in drafting the article, revising article critically for important intellectual content, final approval of the version to be submitted.

Matthew J Grainge: the conception and design of the study, interpretation of data, revising article critically for important intellectual content, final approval of the version to be submitted.

Tim Card: the conception and design of the study, interpretation of data, revising article critically for important intellectual content, final approval of the version to be submitted.

Maarten W Taal: the conception and design of the study, interpretation of data, revising article critically for important intellectual content, final approval of the version to be submitted.

Guruprasad P Aithal: the conception and design of the study, interpretation of data, revising article critically for important intellectual content, final approval of the version to be submitted.

Christopher P Fox: the conception and design of the study, interpretation of data, revising article critically for important intellectual content, final approval of the version to be submitted.

Christian D Mallen: the conception and design of the study, interpretation of data, revising article critically for important intellectual content, final approval of the version to be submitted.

Matthew D Stevenson: the conception and design of the study, interpretation of data, revising article critically for important intellectual content, final approval of the version to be submitted.

Richard D Riley: the conception and design of the study, analysis and interpretation of data, revising article critically for important intellectual content, final approval of the version to be submitted.

**Conflict of interest statement:**

A.A. has received Institutional research grants from AstraZeneca and Oxford Immunotech; and personal fees from UpToDate (royalty), Springer (royalty), Cadilla Pharmaceuticals (lecture fees), NGM Bio (consulting), Limbic (consulting) and personal fees from Inflazome (consulting) unrelated to the work. GP Aithal has received consulting fees from Abbott, Albereo, Amryth, AstraZeneca, BenevolentAI, DNDI, GlaxoSmithKline, NuCANA, Pfizer, Roche Diagnostics, Servier Pharmaceuticals, W.L Gore & Associates paid to the University of Nottingham unrelated to the work. CPF has received Consultancy/Advisory board fees from Abbvie, GenMab, Incyte, Morphosys, Roche, Takeda, Ono, Kite/Gilead, BMS/Celgene, BTG/Veriton and departmental research funding from BeiGene unrelated to the work. The other authors have no conflict of interest to declare.

**Data availability statement:**

Data used in the study are from the Clinical Practice Research Datalink and cannot be shared due to licencing restrictions. Study protocol is available from www.cprd.com.

### Box 1

Equation to predict the risk of 5-ASA discontinuation after six months of primary care prescription and within the next 5-years.

Risk score = 1 – 0.971 ^exp(0.83βX)^, where βX=0.0016945* Age in years at first primary-care prescription + 0.0082816*female-sex + 0.0020271*body mass index - 0.0106059*low alcohol intake + 0.0982098*moderate alcohol intake + 0.6161676*hazardous alcohol intake + 0.4275329 *Ex-alcohol intake + 0.1917626*Psoriasis + 0.2380761*autoimmune rheumatic disease + 0.1383201*diabetes + 0.8360309*Chronic kidney disease stage-3 + 0.7359997*thiopurine + 0.6131115*methotrexate or leflunomide + 0.1890031*statins + 0.2554022*Carbamazepine/valproate + 0.231411*paracetamol + 0.4665026* At-least mild cytopenia or liver enzyme elevation within first six-months of primary-care prescription.

All variables are code 0, and 1 if absent or present respectively, except for BMI and age that are continuous variables. 0.971 is the baseline survival function at 5-years, 0.83 is the shrinkage factor and the other numbers are the estimated regression coefficients for the predictors, which indicate their mutually adjusted relative contribution to the outcome risk. Thiopurines included either azathioprine or 6-mercaptopurine. Autoimmune rheumatic diseases included rheumatoid arthritis, systemic lupus erythematosus, ankylosing spondylitis, reactive arthritis.

## Data Availability

Data used in the study are from the Clinical Practice Research Datalink and cannot be shared due to licensing restrictions. Study protocol is available from www.cprd.com.

## Funding

This research was funded by National Institute for Health and Care Research (NIHR) grant NIHR130580. The funders had no role in conducting and/or reporting this study.

---

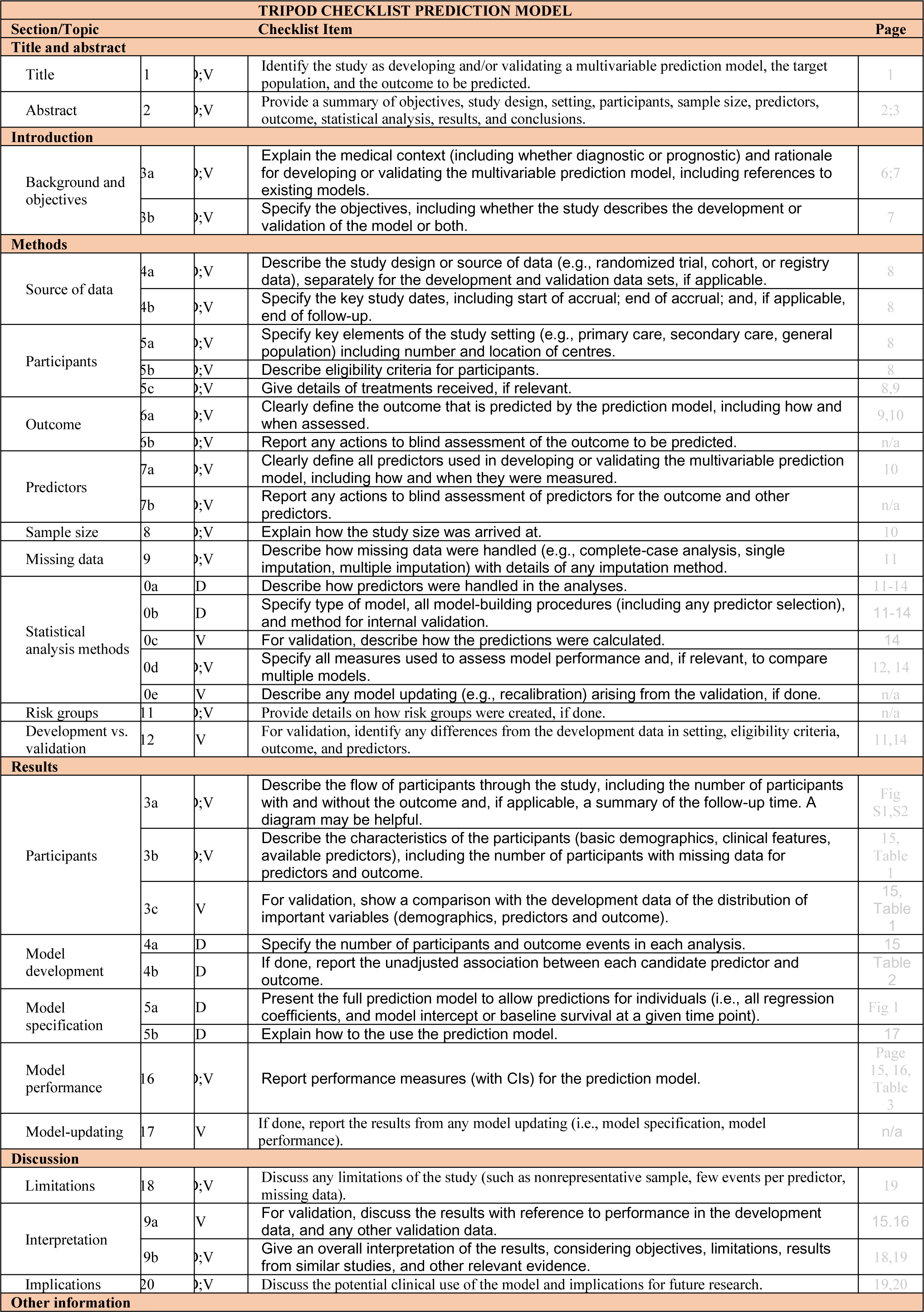
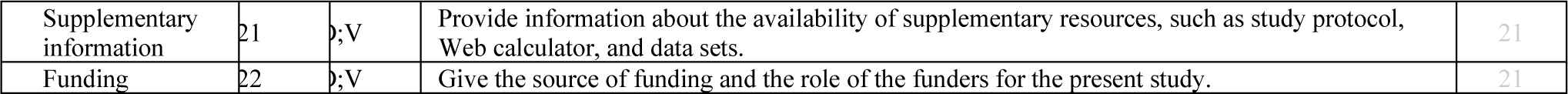

## Notes

### Author Declarations

This study was approved through the Clinical Practice Research Datalink (CPRD)'s Research Data Governance process (Reference: 20_000236R).

