## supplementary material for "Monitoring for 5-aminosalicylate toxicity: prognostic model development and validation"

### Contents

Supplementary Table 1: Individual patient's characteristics at the midpoint of each decile. 2

Supplementary Table 2: The estimated probability that a delayed abnormal blood test would have caused a more serious condition with 5-ASA. 3

Supplementary Table 3: The estimated costs and Quality Adjusted Life Years (QALY) losses associated with each condition derived from the literature[1-11]. Please see health economics methods. 4

Supplementary Table 4: Disaggregated results in the base case 5

Supplementary Table 5: Aggregated Results in the base case compared with current 3-month monitoring 5

Figure S1: Study population selection criteria for model development 6

Figure S2: Model validation cohort: Study population selection criteria 7

Figure S3: Nelson–Aalen cumulative hazard estimates for 5-ASA discontinuation associated with individual blood-test abnormalities* 8

Figure S4: Distribution of predicted risk in the model development cohort at 5 years 9

Figure S5: Calibration plot of a prognostic model for 5-ASA discontinuation with abnormal monitoring blood-test results at 5-years in the development cohort[1] 10

Figure S6: Calibration plot of a prognostic model for 5-ASA discontinuation with abnormal monitoring blood-test results at 5-years in the validation cohort[1] 11

Figure S7: Distribution of predicted risk in the model validation cohort at 5 years 12

Figure S8: Calibration of a prognostic model for 5-ASA discontinuation with abnormal monitoring blood-test results at 1 year in the validation cohort[1] 13

Figure S9: Calibration of a prognostic model for 5-ASA discontinuation with abnormal monitoring blood-test results at 2 years in the validation cohort[1] 14

Figure S10: Calibration of a prognostic model for 5-ASA discontinuation with abnormal monitoring blood-test results at 3 years in the validation cohort[1] 15

Figure S11: Calibration of a prognostic model for 5-ASA discontinuation with abnormal monitoring blood-test results at 4 years in the validation cohort[1] 16

Figure S12: Incremental net monetary benefit results when the risks of serious conditions estimated by clinicians were tripled 17

#### Supplementary Table 1: Individual patient's characteristics at the midpoint of each decile.

| Decile | Age (yr.) | Sex | BMI (kg/m$^2$) | Alcohol | Disease | DM | CKD -3 | IS drug | Statins | Carb/ Val | Paracetamol | BTA | Cumulative probability of outcome % |
|---|---|---|---|---|---|---|---|---|---|---|---|---|---|
| 1 | 30-35 | M | 24.6 | Low | IBD | No | No | No | No | No | No | No | 3.13 |
| 2 | 35-40 | M | 23.8 | Low | IBD | No | No | No | No | No | No | No | 3.18 |
| 3 | 30-35 | F | 28.0 | Low | IBD | No | No | No | No | No | No | No | 3.23 |
| 4 | 50-55 | F | 19.1 | Non-drinker | IBD | No | No | No | No | No | No | No | 3.29 |
| 5 | 65-70 | F | 28.5 | Low | IBD | No | No | No | No | No | Yes | No | 3.39 |
| 6 | 65-70 | M | 26.1 | Low | IBD | No | No | No | Yes | No | Yes | No | 3.88 |
| 7 | 50-55 | M | 27.9 | Non-drinker | IBD | No | No | No | No | No | No | No | 4.55 |
| 8 | 40-45 | M | 22.9 | Non-drinker | IBD | Yes | No | No | No | No | Yes | No | 5.48 |
| 9 | 65-70 | M | 31.0 | Low | IBD | No | Yes | Aza/6-MP | No | No | Yes | No | 6.08 |
| 10 | 60-65 | M | 30.7 | Low | IBD | Yes | Yes | No | Yes | No | No | No | 8.62 |

Aza/6-MP: Azathioprine/6-Mercaptopurine; BMI: Body Mass Index; BTA: Blood Test abnormalities within 6 months of primary care 5-ASA prescription; CKD-3: Chronic Kidney Disease stage 3; Carb/Val: carbamazepine/valproate; DM: diabetes mellitus; F: female; IS immunosuppressive; M: male, ± 5-year age band. Exact age not shown for anonymity.

Supplementary Table 2: The estimated probability that a delayed abnormal blood test would have caused a more serious condition with 5-ASA.

| Adverse Event | 6-monthly monitoring | Annual monitoring | Two-yearly monitoring |
|---|---|---|---|
| Acute kidney injury* | | | |
| Acute liver failure* | | | |
| Anaemia* | | | |
| Chronic kidney disease | 0.001 | 0.001 | 0.008 |
| Cirrhosis* | | | |
| Drug-induced liver injury | 0.001 | 0.001 | 0.001 |
| Early fibrosis* | | | |
| Low neutrophil count plus sepsis | 0.0001 | 0.0002 | 0.0004 |
| Neutropenic sepsis | 0.0001 | 0.0002 | 0.0004 |
| Thrombocytopenia requiring hospitalisation* | | | |
| Thrombocytopenia with superficial bleeding* | | | |

*The clinical experts did not think that 5-ASA causes these outcomes.

Supplementary Table 3: The estimated costs and Quality Adjusted Life Years (QALY) losses associated with each condition derived from the literature[1-11]. Please see health economics methods.

| Condition | Costs (£) | QALY loss |
| --- | --- | --- |
| Acute Kidney Injury | 2022 | 0.014 |
| Acute Liver Failure | 3352 | 0.651 |
| Anaemia | 465 | 0.001 |
| Chronic Kidney Disease | 26,083 | 1.308 |
| Cirrhosis | 21,700 | 3.042 |
| Drug-Induced Liver Injury | 3352 | 0.651 |
| Early Fibrosis | 0 | 0.075† |
| Low neutrophil count plus sepsis | 2313 | 2.490 |
| Neutropenic Sepsis | 9456 | 2.490 |
| Thrombocytopenia requiring hospitalisation | 1018 | 0.016 |
| Thrombocytopenia with superficial bleeding | 133 | 0.001 |

†per 6 months unidentified

#### Supplementary Table 4: Disaggregated results in the base case

| | Monitoring Appointments | | | | Monitoring Costs Saved compared with current 3-month monitoring (£) | | | Abnormal blood results identified late compared with current 3-month monitoring | | | Costs associated with late identification of abnormal blood results (£) | | | QALY losses associated with late identification of abnormal blood results | | |
|---|---|---|---|---|---|---|---|---|---|---|---|---|---|---|---|---|
| Decile | 3-months | 6-month | Annual | Biennial | 6-month | Annual | Biennial | 6-month | Annual | Biennial | 6-month | Annual | Biennial | 6-month | Annual | Biennial |
| 1 | 19.67 | 9.84 | 4.93 | 1.99 | 185.99 | 278.98 | 331.62 | 0.016 | 0.024 | 0.029 | 0.46 | 0.73 | 5.94 | 0.0000 | 0.0001 | 0.0004 |
| 2 | 19.67 | 9.84 | 4.93 | 1.99 | 185.98 | 278.96 | 331.59 | 0.016 | 0.025 | 0.029 | 0.46 | 0.73 | 5.96 | 0.0000 | 0.0001 | 0.0004 |
| 3 | 19.67 | 9.84 | 4.93 | 1.99 | 185.95 | 278.92 | 331.54 | 0.016 | 0.025 | 0.029 | 0.47 | 0.74 | 6.01 | 0.0000 | 0.0001 | 0.0004 |
| 4 | 19.66 | 9.84 | 4.93 | 1.99 | 185.88 | 278.81 | 331.42 | 0.017 | 0.025 | 0.030 | 0.48 | 0.76 | 6.14 | 0.0000 | 0.0001 | 0.0004 |
| 5 | 19.65 | 9.83 | 4.93 | 1.99 | 185.80 | 278.69 | 331.28 | 0.017 | 0.026 | 0.031 | 0.49 | 0.77 | 6.29 | 0.0000 | 0.0001 | 0.0004 |
| 6 | 19.60 | 9.81 | 4.91 | 1.99 | 185.28 | 277.91 | 330.34 | 0.020 | 0.030 | 0.036 | 0.56 | 0.89 | 7.26 | 0.0000 | 0.0001 | 0.0004 |
| 7 | 19.52 | 9.77 | 4.90 | 1.99 | 184.48 | 276.70 | 328.90 | 0.024 | 0.036 | 0.043 | 0.68 | 1.08 | 8.74 | 0.0001 | 0.0001 | 0.0005 |
| 8 | 19.45 | 9.74 | 4.88 | 1.98 | 183.82 | 275.71 | 327.71 | 0.027 | 0.043 | 0.049 | 0.77 | 1.23 | 9.97 | 0.0001 | 0.0001 | 0.0006 |
| 9 | 19.36 | 9.70 | 4.86 | 1.98 | 183.01 | 274.50 | 326.26 | 0.031 | 0.047 | 0.056 | 0.89 | 1.41 | 11.46 | 0.0001 | 0.0001 | 0.0007 |
| 10 | 19.11 | 9.58 | 4.81 | 1.97 | 180.54 | 270.79 | 321.82 | 0.043 | 0.066 | 0.080 | 1.24 | 1.97 | 16.00 | 0.0001 | 0.0002 | 0.0010 |

#### Supplementary Table 5: Aggregated Results in the base case compared with current 3-month monitoring

| | Net Cost Savings (£) | | | Net QALY losses | | |
|---|---|---|---|---|---|---|
| Decile | 6-month | Annual | Biennial | 6-month | Annual | Biennial |
| 1 | 185.53 | 278.25 | 325.68 | 0.0000 | 0.0001 | 0.0004 |
| 2 | 185.51 | 278.22 | 325.63 | 0.0000 | 0.0001 | 0.0004 |
| 3 | 185.48 | 278.18 | 325.53 | 0.0000 | 0.0001 | 0.0004 |
| 4 | 185.40 | 278.05 | 325.27 | 0.0000 | 0.0001 | 0.0004 |
| 5 | 185.31 | 277.92 | 324.99 | 0.0000 | 0.0001 | 0.0004 |
| 6 | 184.71 | 277.02 | 323.08 | 0.0000 | 0.0001 | 0.0004 |
| 7 | 183.80 | 275.63 | 320.16 | 0.0001 | 0.0001 | 0.0005 |
| 8 | 183.04 | 274.49 | 317.75 | 0.0001 | 0.0001 | 0.0006 |
| 9 | 182.12 | 273.09 | 314.80 | 0.0001 | 0.0001 | 0.0007 |
| 10 | 179.29 | 268.82 | 305.83 | 0.0001 | 0.0002 | 0.0010 |

#### Figure S1: Study population selection criteria for model development

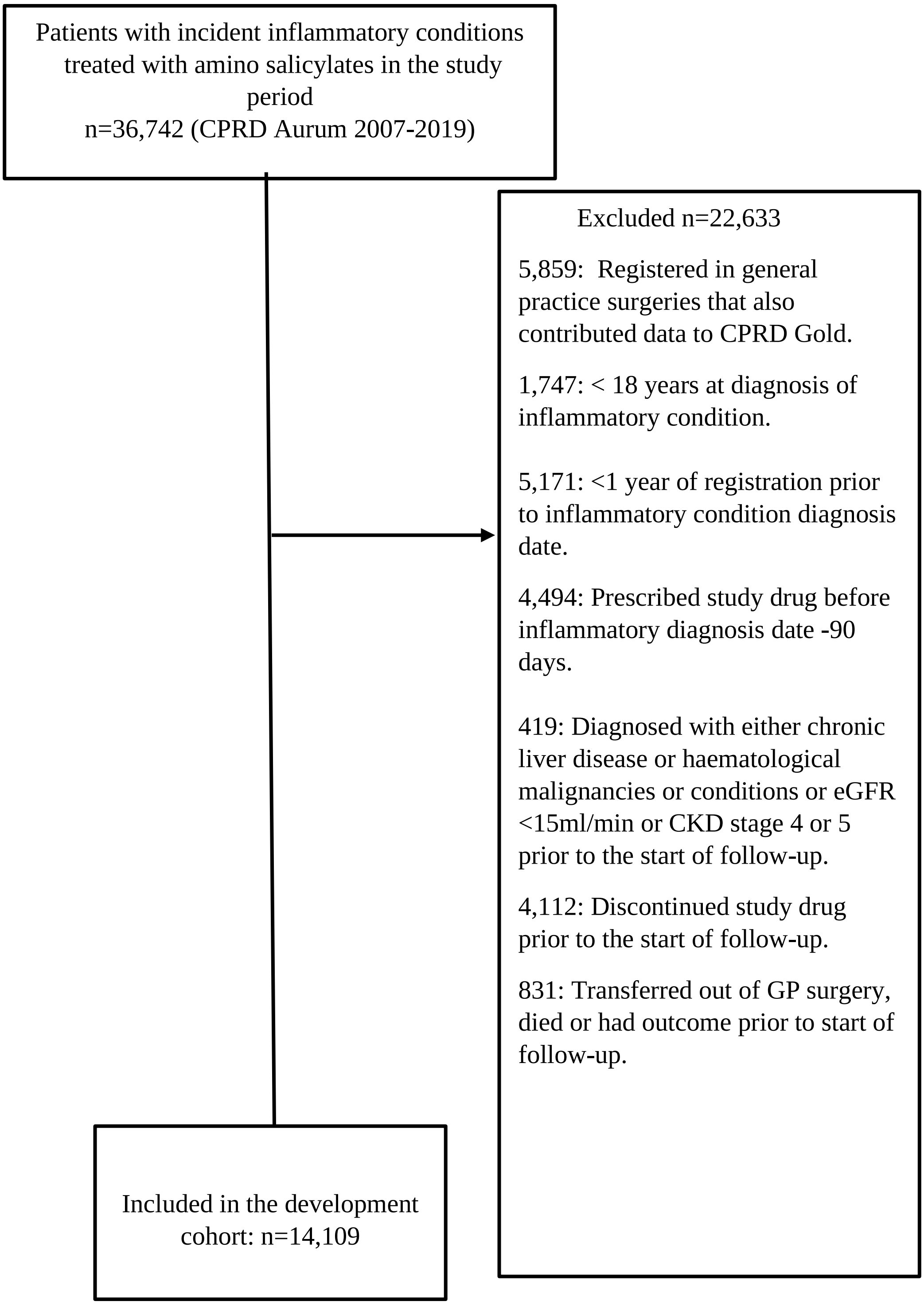


Aminosalicylates were balsalazide, mesalazine and olsalazine, CKD: chronic kidney disease, GP: General practice, CPRD: Clinical Practice Research Datalink.

#### Figure S2: Model validation cohort: Study population selection criteria

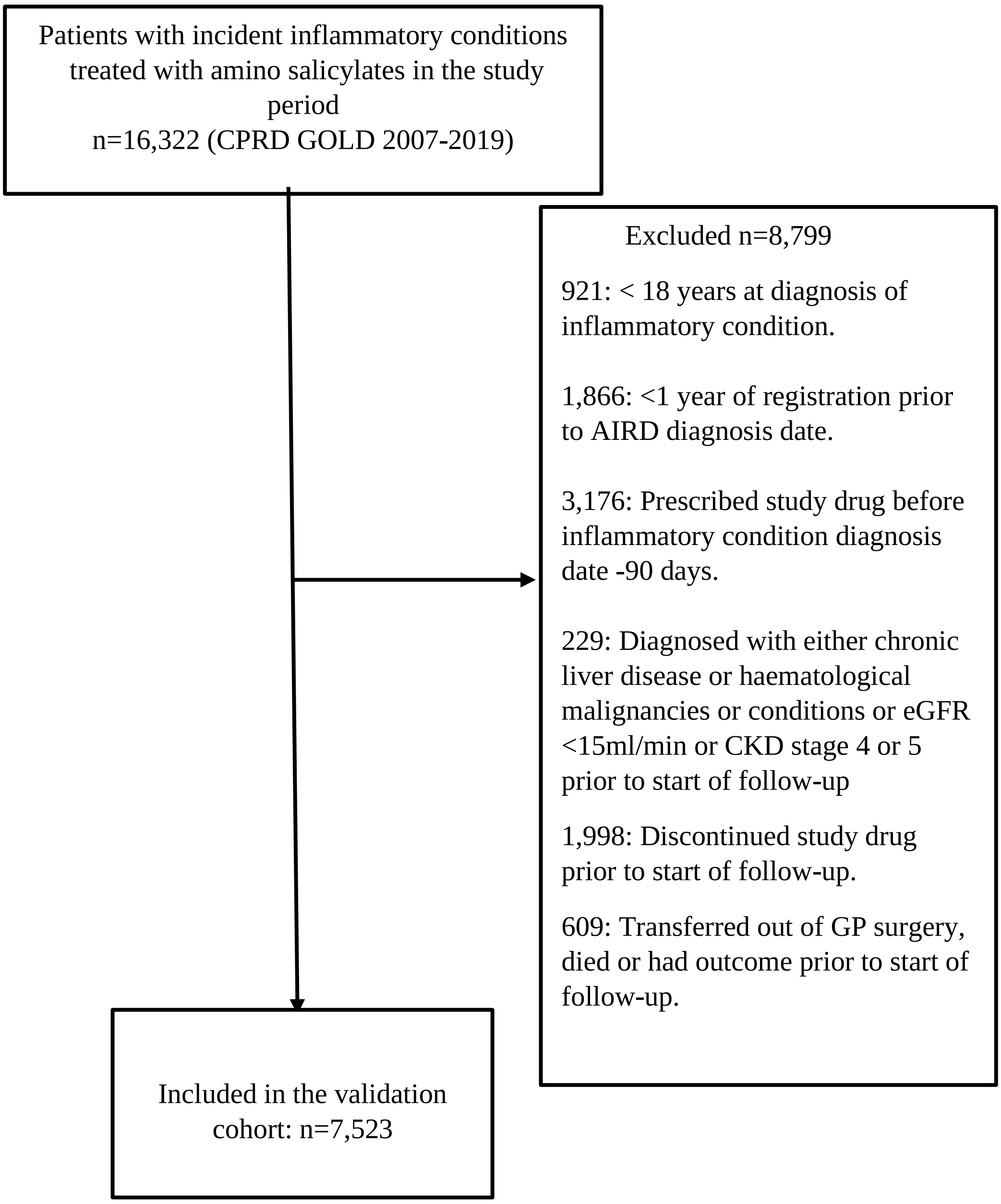


Aminosalicylates were balsalazide, mesalazine and Olsalazine, CKD: chronic kidney disease, GP: General practice, CPRD: Clinical Practice Research Datalink.

Figure S3: Nelson–Aalen cumulative hazard estimates for 5-ASA discontinuation associated with individual blood-test abnormalities*.

A.

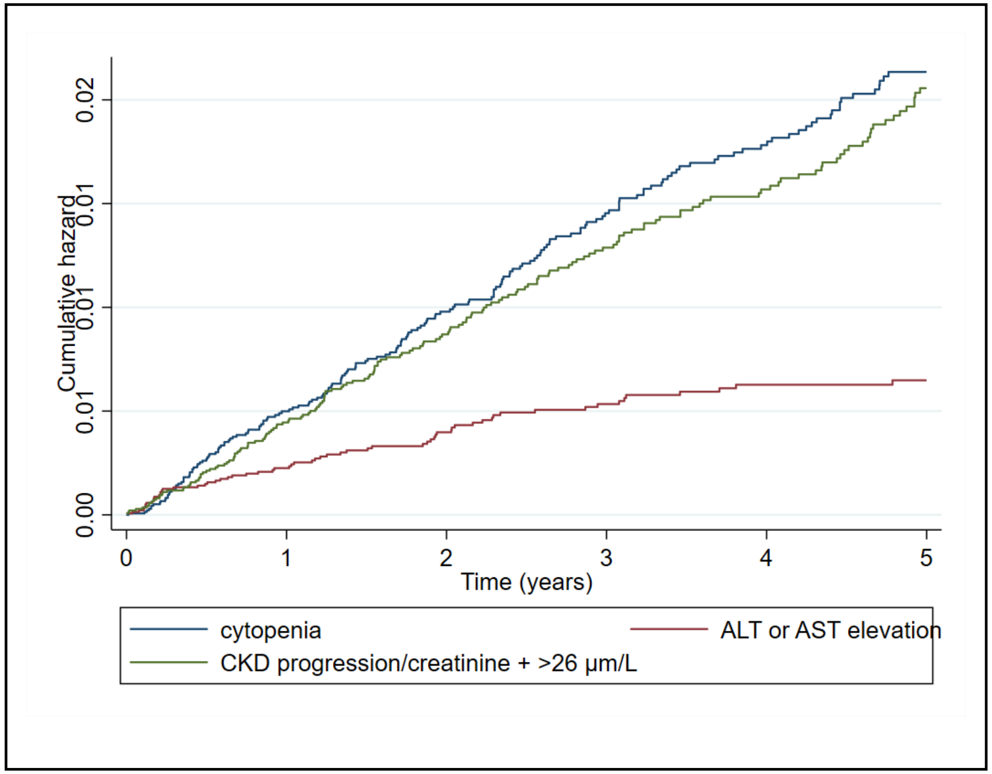


B

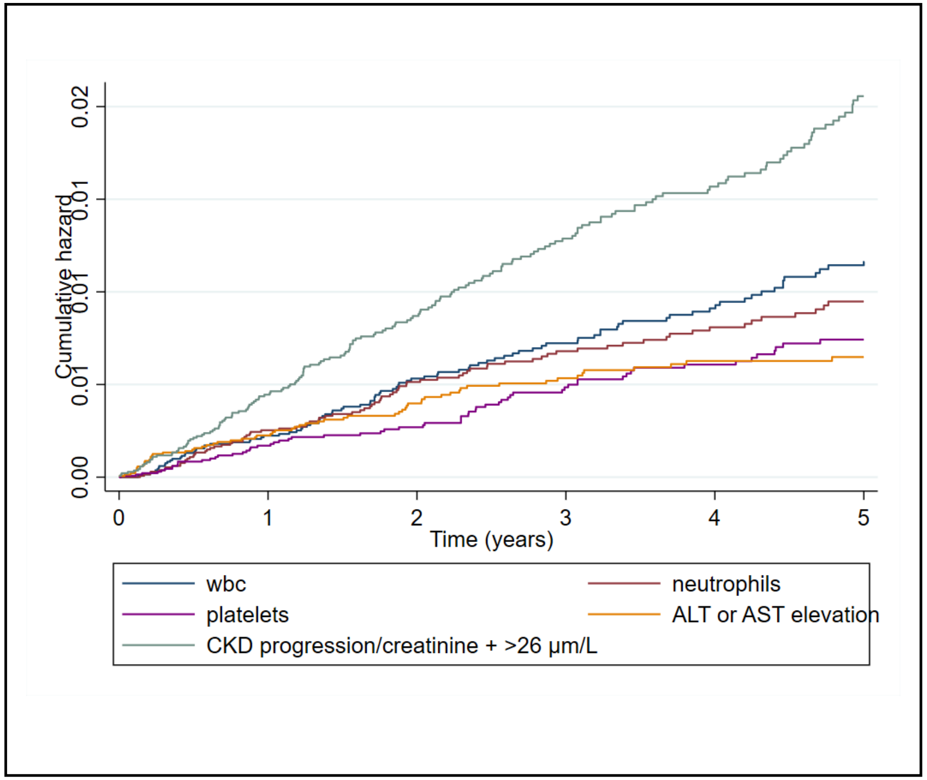


* cytopenia defined as white blood cells (WBCs) <3.5 × $10^9$/l or neutrophils <1.6 × $10^9$/l or platelets <140 × $10^9$/l; ALT/AST >100 IU/l

Figure S4: Distribution of predicted risk in the model development cohort at 5 years

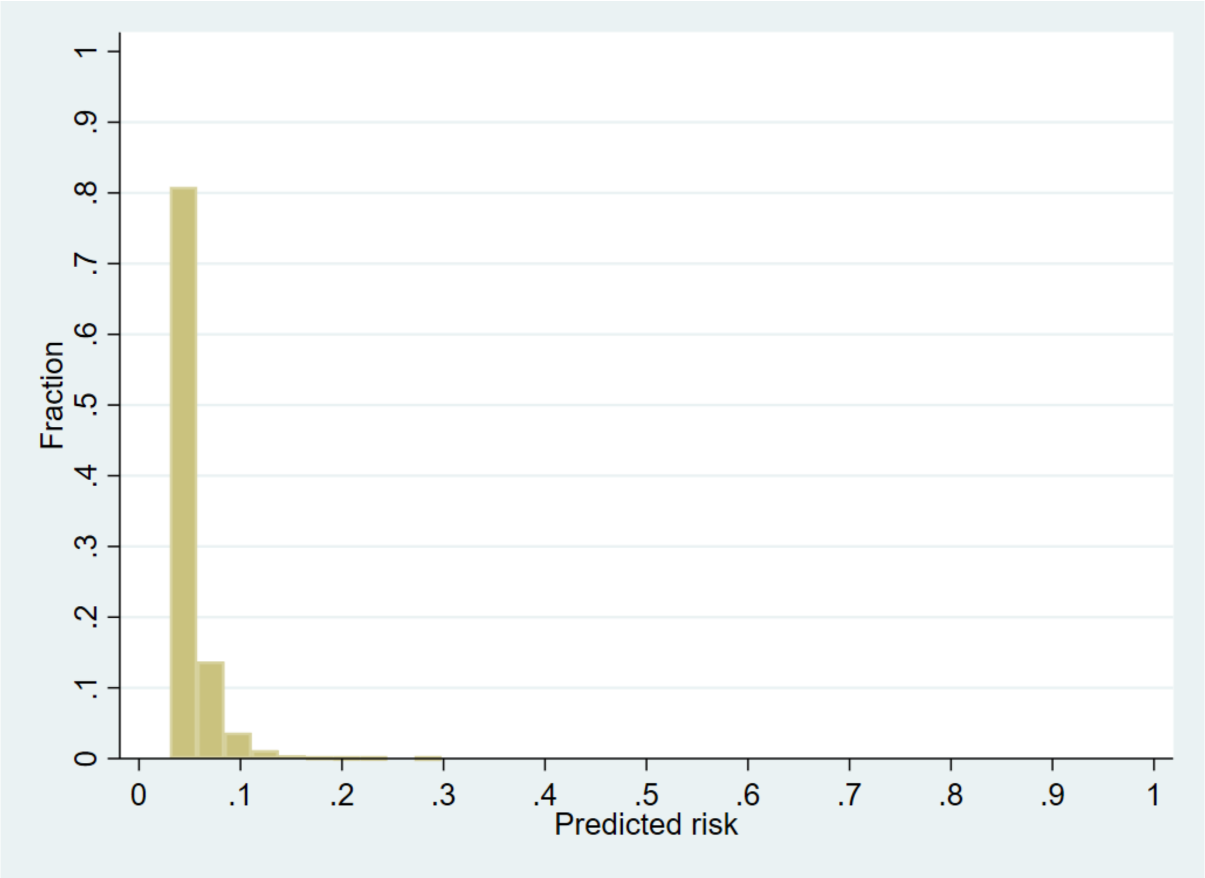

Figure S5: Calibration plot of a prognostic model for 5-ASA discontinuation with abnormal monitoring blood-test results at 5-years in the development cohort[1]

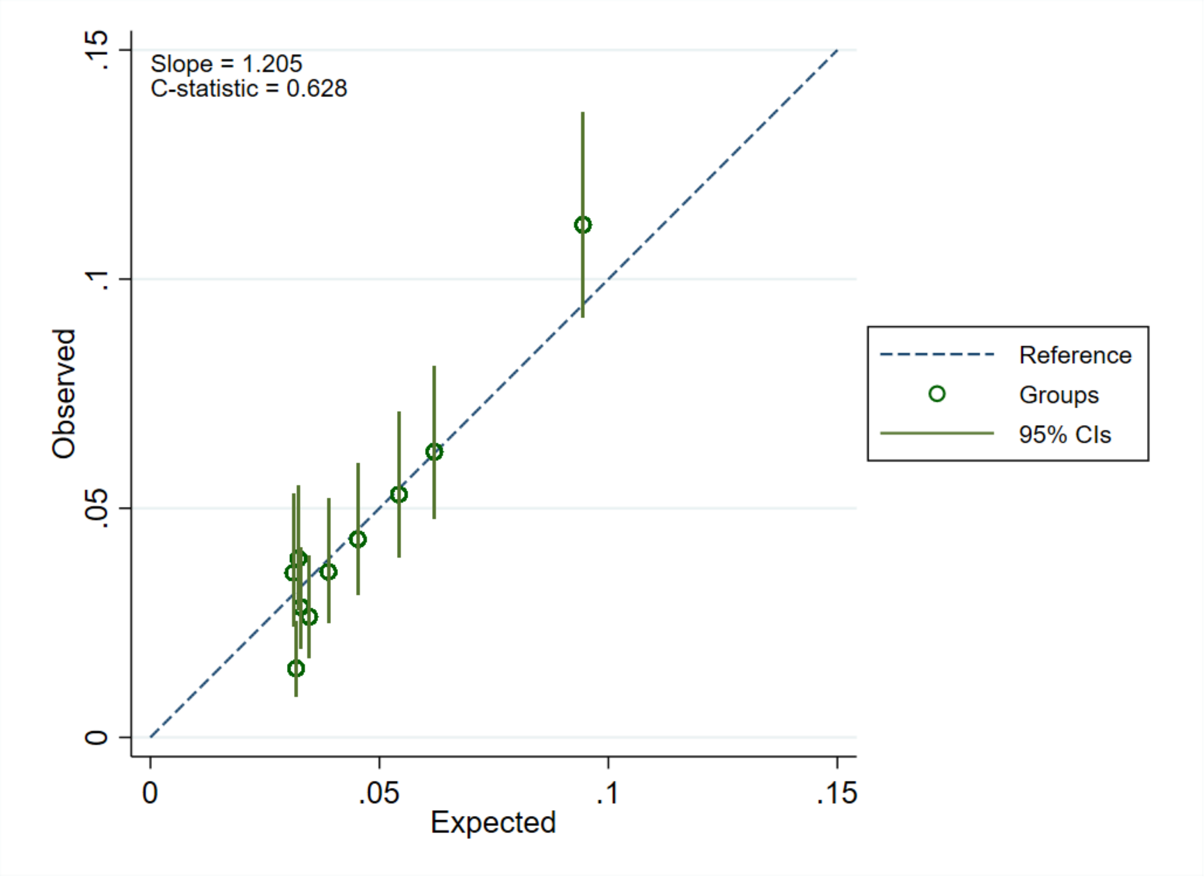


[1]Data from a single imputed dataset; So(t=5) =0.971

Figure S6: Calibration plot of a prognostic model for 5-ASA discontinuation with abnormal monitoring blood-test results at 5-years in the validation cohort[1]

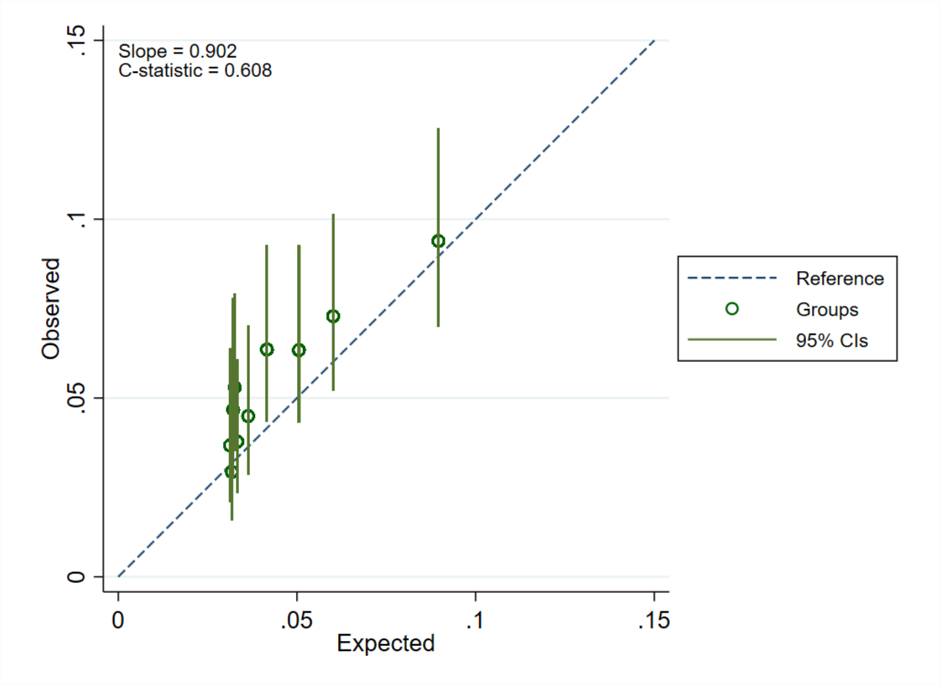


[1]Data from a single imputed dataset; So(t=5) =0.971

Figure S7: Distribution of predicted risk in the model validation cohort at 5 years

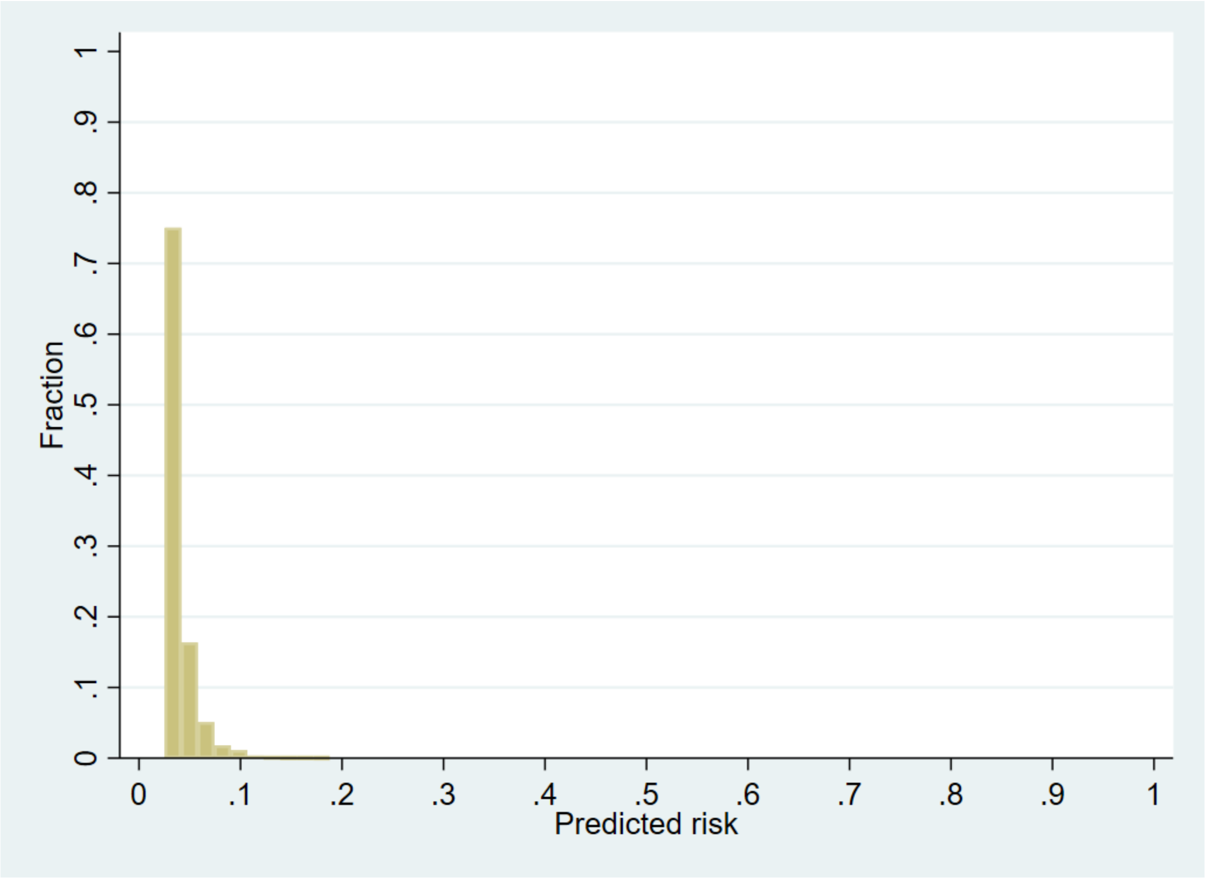

Figure S8: Calibration of a prognostic model for 5-ASA discontinuation with abnormal monitoring blood-test results at 1 year in the validation cohort[1]

A: Calibration plot

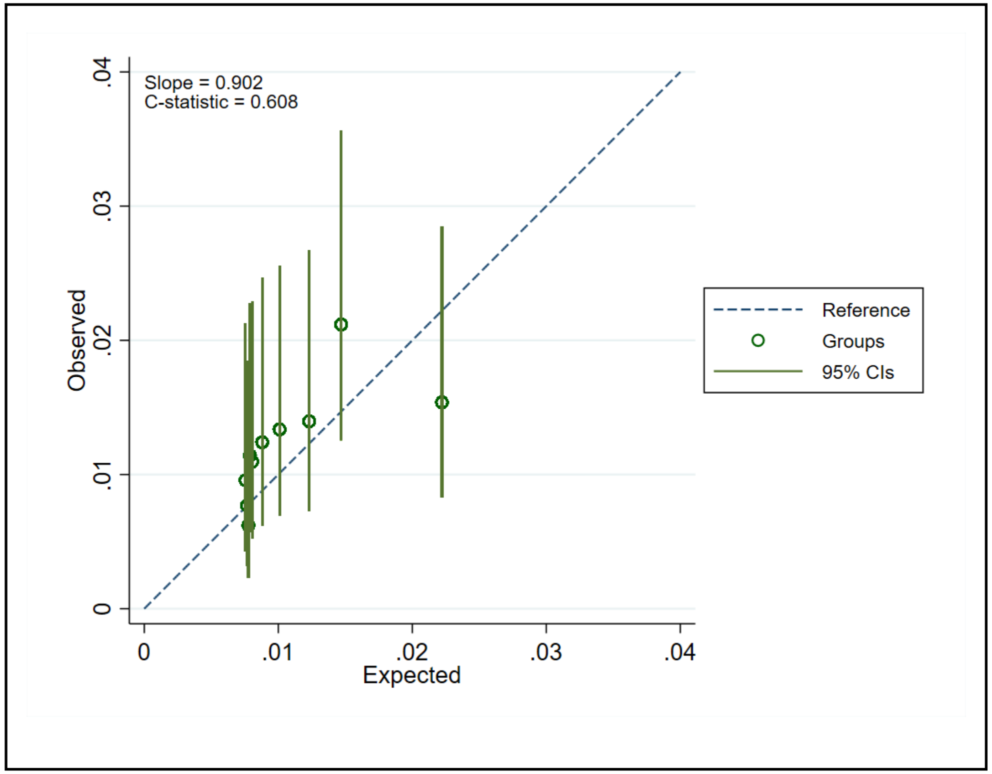


B: Smoothed calibration curve

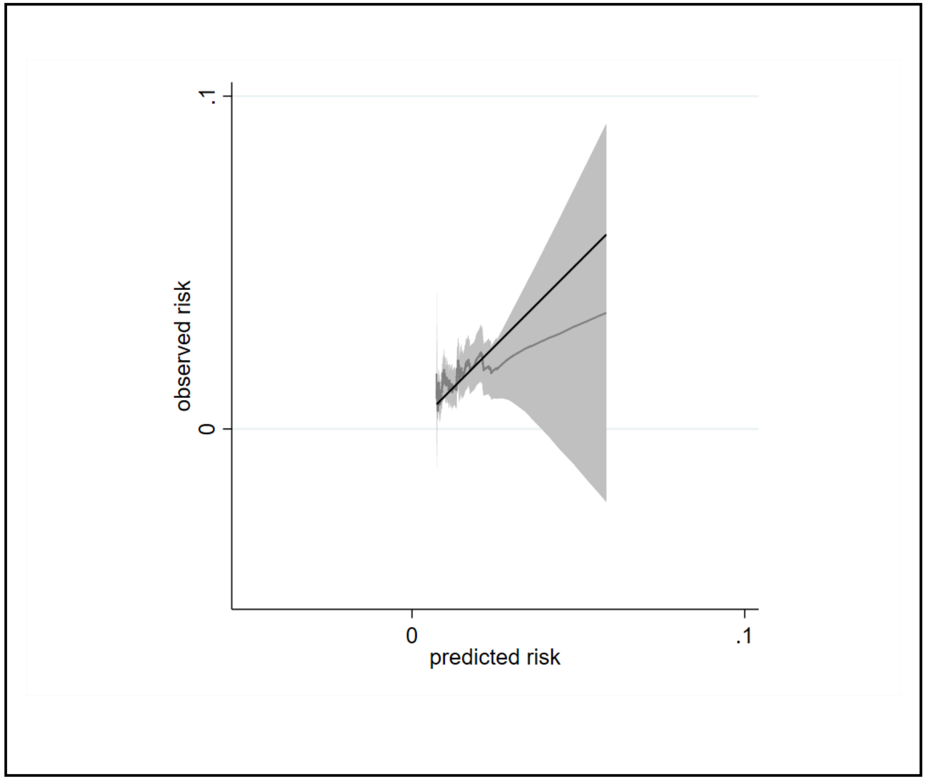


[1]Data from a single imputed dataset was used; $S_0(t_{=1})$ =0.993

Figure S9: Calibration of a prognostic model for 5-ASA discontinuation with abnormal monitoring blood-test results at 2 years in the validation cohort[1]

A. Calibration plot

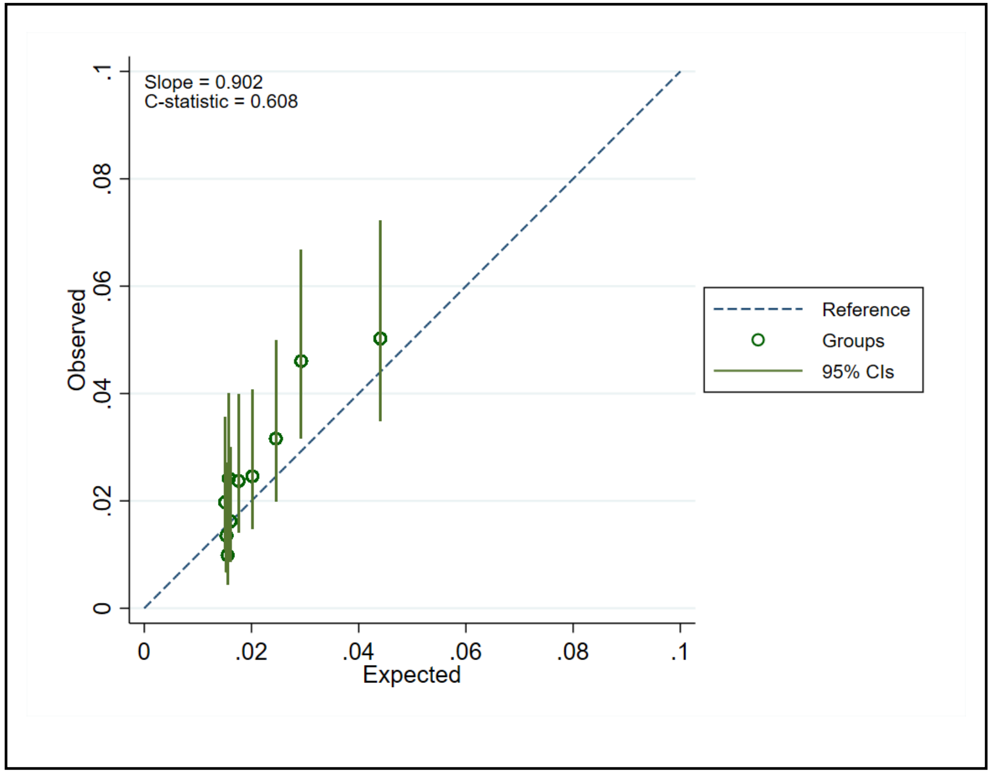


B. Smoothed calibration curve

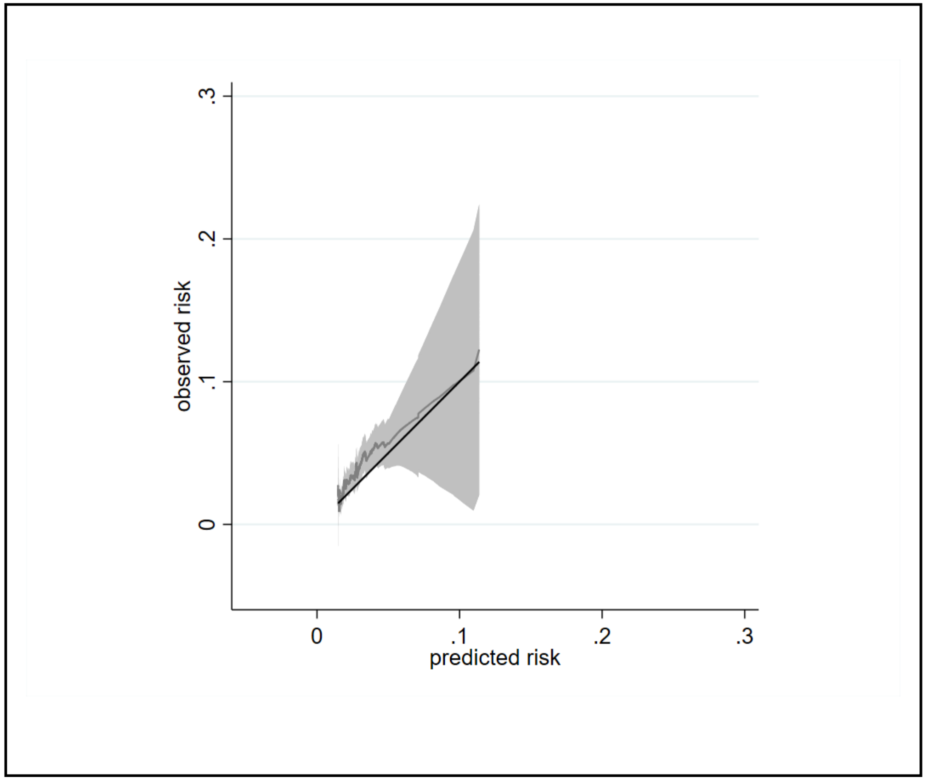


[1]Data from a single imputed dataset was used; $S_0(t_{=2})$ =0.986

Figure S10: Calibration of a prognostic model for 5-ASA discontinuation with abnormal monitoring blood-test results at 3 years in the validation cohort[1]

A. Calibration plot

B. Smoothed calibration curve

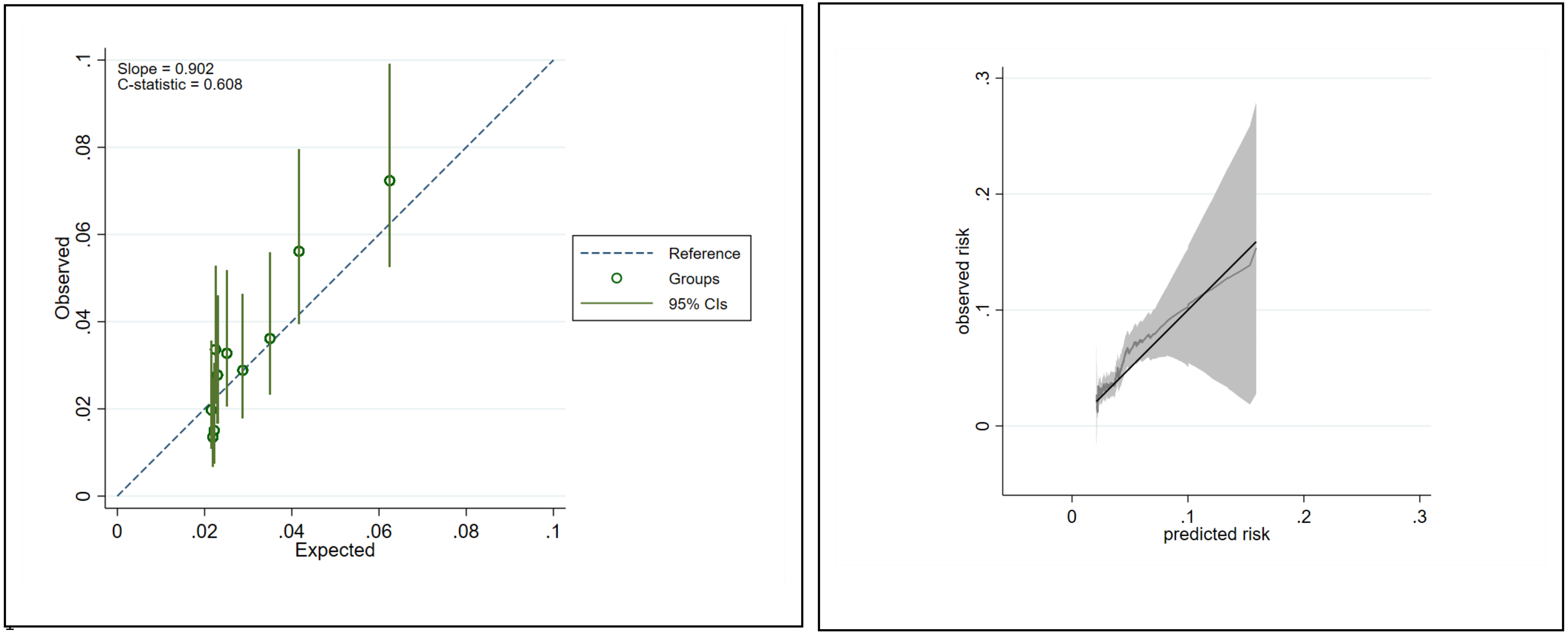


Data from a single imputed dataset was used; $S_0(t_{=3})$ =0.980

Figure S11: Calibration of a prognostic model for 5-ASA discontinuation with abnormal monitoring blood-test results at 4 years in the validation cohort[1]

A. Calibration plot

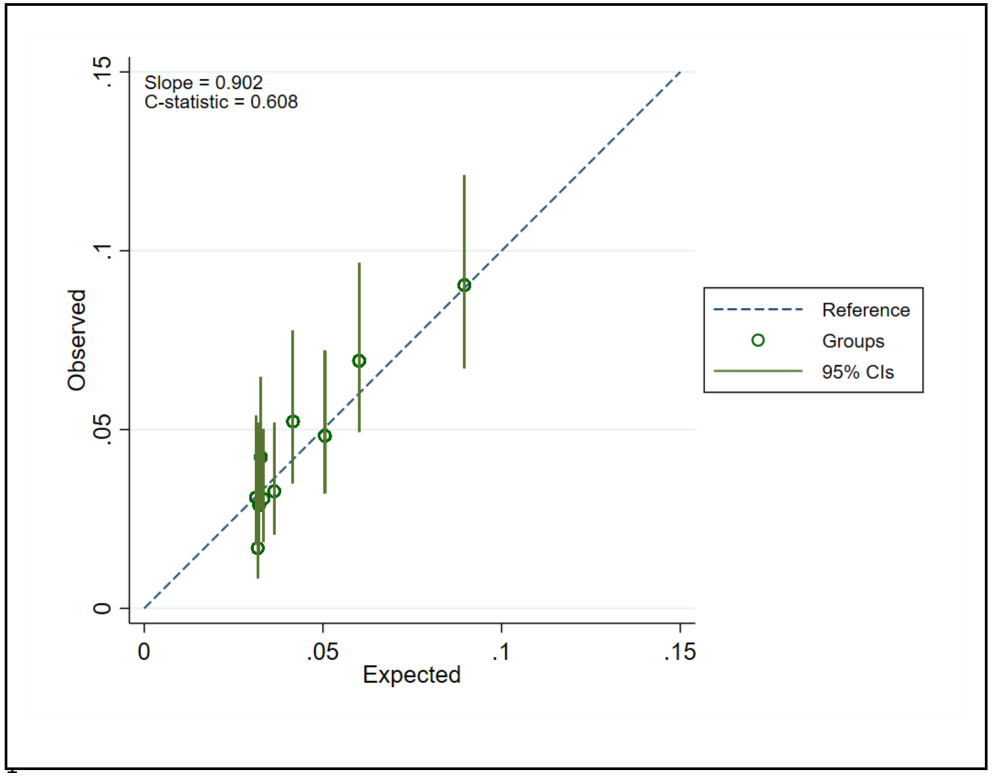


B. Smoothed calibration curve

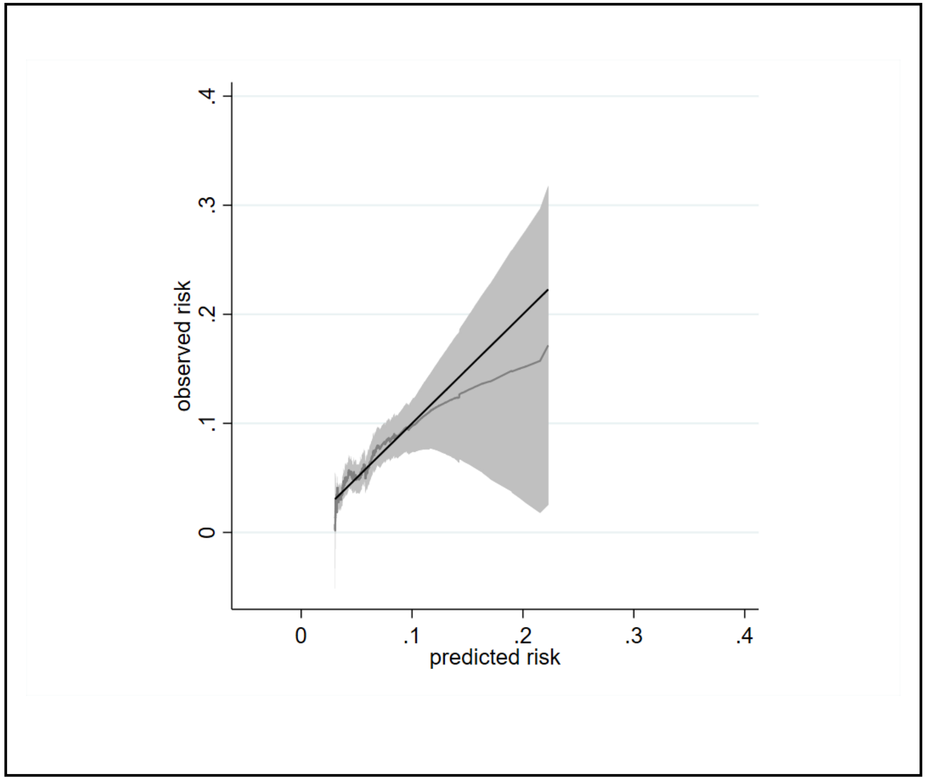


Data from a single imputed dataset was used; $S_0(t_{=4})$=0.976

Figure S12: Incremental net monetary benefit results when the risks of serious conditions estimated by clinicians were tripled

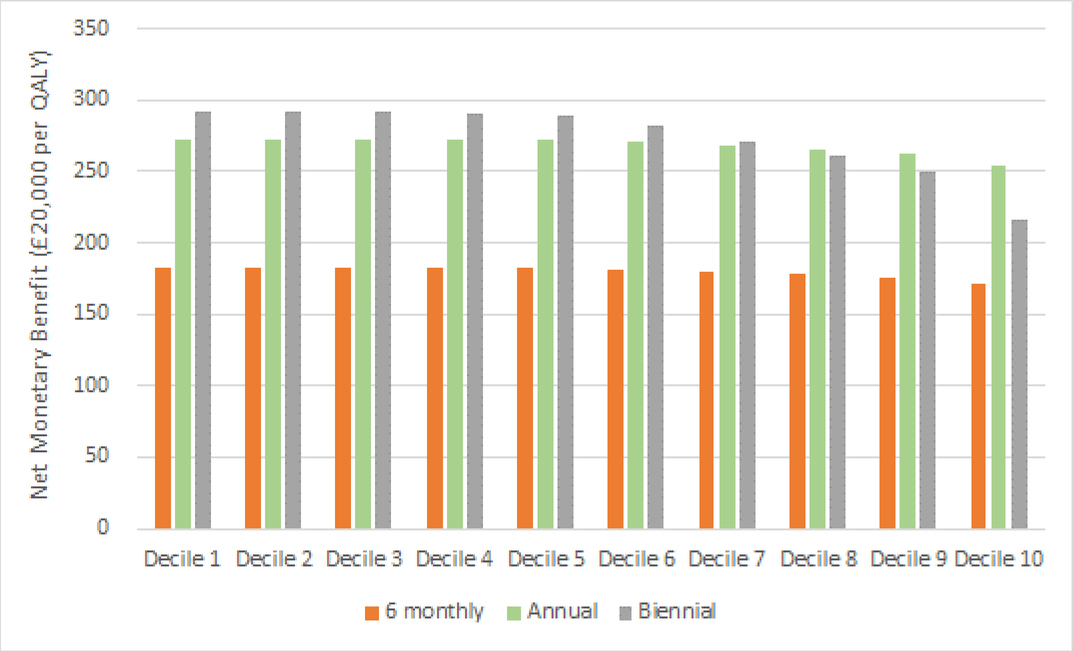
